# Medical cannabis in the UK: prescriptions, sources, products, and high-risk use

**DOI:** 10.64898/2026.01.19.26344380

**Authors:** Elle Wadsworth, David Hammond, Tom P Freeman

## Abstract

**Background and Aims:** The UK legalised medical cannabis in 2018, yet little is known about people accessing cannabis legally via prescription or those using cannabis for medical purposes without a prescription. This study aimed to estimate: 1) the percentage of people who use cannabis medically with and without a prescription; 2) the sources used to obtain cannabis; 3) the products used; and 4) the associations between monthly or more frequent (MMF) product use and medical cannabis use status.

**Design:** National repeat cross-sectional surveys conducted in September-November 2023 and 2024.

**Setting:** UK.

**Participants:** People aged 16-65 who used cannabis in the past 12 months (n=4,414).

**Measurements:** Inferential statistics compared outcomes by medical cannabis use status. Multivariable regression analyses estimated associations between MMF product use and medical cannabis use status.

**Findings:** Overall, 13.0% of people reported receiving a medical cannabis prescription, 36.4% reported medical use without a prescription, and 50.6% reported no medical use. Cannabis was sourced through diverse routes; only 10.7% of people with prescriptions obtained all their cannabis from a legal medical prescription. People with a medical prescription had a higher probability of reporting MMF use of oils or liquid drops (aRR=3.51; 95% CI: 2.73-4.52), oil or liquid capsules (aRR=2.63; 1.99-3.47), vape oils (aRR=2.40; 2.03-2.84), edibles (aRR=2.41; 2.01-2.90), cannabis drinks (aRR=3.39; 2.69-4.26), solid concentrates (aRR=3.10; 2.29-4.21), hash or kief (aRR=1.82; 1.44-2.30), and topicals (aRR=3.42; 2.58-4.51) than people who did not report medical use, after adjusting for covariates. People who ever asked for a medical prescription and people with a medical prescription in the past year had a higher probability of screening positive for high-risk use.

**Conclusion:** Over one in ten people who use cannabis in the UK report a medical cannabis prescription, yet a third reported medical use without a prescription. With only a minority obtaining all their cannabis legally, many rely on illegal sources, suggesting prescriptions may not meet the needs of those reporting use for medical purposes. People with prescriptions show a higher probability of frequent use of processed, higher-potency products, and meeting a threshold for high-risk use. Healthcare encounters during medical cannabis prescribing should discuss risks related to potency, product type, and adverse effects such as cannabis use disorder.

## INTRODUCTION

Cannabis is the most used illegal drug in the UK, with 7.6% and 6.8% of adults in England and Wales reporting past year use in 2023 and 2024 (1). In addition to using cannabis for nonmedical (or “recreational”) purposes, cannabis can be used for therapeutic benefits.

Evidence for the effectiveness of therapeutic benefits of cannabis, both for physical and mental health conditions, is limited (2, 3). Regardless, cannabis is still widely used for potential therapeutic benefits, such as pain relief (4–8), with the more substantial evidence of effectiveness pointing to neuropathic pain, cancer pain, chemotherapy-induced nausea and multiple sclerosis spasticity symptoms (9). In cross-sectional surveys conducted among UK adults, consumers using cannabis for medical reasons were most likely to report using cannabis to manage depression, anxiety and chronic pain (10, 11).

In recent years, medical and nonmedical cannabis policies have become more permissive globally. In the UK, nonmedical cannabis is illegal; however, unapproved/unlicensed cannabis-based products for medical use (CBPM) became legal to prescribe in November 2018. Medical cannabis prescriptions can be obtained from specialist doctors registered with the General Medical Council in either the National Health Service (NHS) or through private cannabis clinics (12). These prescriptions are intended for those who have exhausted all other recommended treatment options. There are barriers to accessing legal medical cannabis: access to medical cannabis prescriptions given through the NHS remains very restrictive, with the majority accessing through private clinics (13). In contrast to the small number of consumers accessing medical cannabis legally, use of ‘illegal’ cannabis for medical purposes is widespread (10, 11). A nationally representative survey estimated that approximately 1.4 million UK residents were consuming cannabis for medical purposes (10). Thus, there seems to be an unmet need of medical cannabis consumers, who are not accessing legally perhaps due to affordability, accessibility, and/or or stigma (14–16). Beyond data collected on patient outcomes from medical cannabis patient registries, little is known about those accessing cannabis legally via prescription, as well as those using cannabis for medical purposes but accessing illegally, including the types of cannabis products accessed and used.

Cannabis contains over 100 cannabinoids, and the primary cannabinoids of interest are Δ9-tetrahydrocannabinol (THC), which is the main psychoactive compound, and cannabidiol (CBD), a non-psychoactive compound (17). The THC concentration of dried flower has been increasing for several decades (18). The increasing level of potency is a public health concern given the association between high-potency products and increased risk of psychotic disorders and cannabis use disorders (19–23). The psychoactive effects of cannabis depend on the type of product (e.g., dried flower, or “processed” products such as extracts), route of administration (e.g., smoked), and potency. Smoking dried flower with tobacco is the most common method of cannabis consumption in the UK; however, evidence suggests that the market is diversifying to more processed (i.e., non-flower) products, which tend to have greater potency (24, 25). It is unclear whether the patterns seen in the broader UK cannabis market is mirrored among those who use cannabis for medical purposes or the implications of such product use. Indeed, not all product forms are permitted in the legal medical market. Permitted forms of unapproved/unlicensed CBPM’s in the UK medical market include dried flower that is recommended to be vaped, oils/tinctures and capsules; other processed products, such as hash, solid concentrates or vape cartridges or edibles (e.g., gummies) are not permitted (2, 26). However, the level of adherence to this guidance is unclear. It is therefore critical to assess the type of products being used by people consuming cannabis for reported medical reasons, to understand health effects in this important subgroup who have not been characterised in previous studies.

The UK Advisory Council for the Misuse of Drugs assessed the impact of rescheduling medical cannabis in 2020 and stated that more data and research were needed (27). In 2025, they issued a call for evidence into cannabis-based products for medical use, requesting information on barriers to access, perceptions and unintended consequences of the policy change (28). As such, the aims of the study were to estimate: 1) the percentage of people who use cannabis for medical purposes and who have and do not have a medical cannabis prescription; 2) the sources used to obtain cannabis; 3) the products used; and 4) associations between product use and receiving a medical cannabis prescription.

## METHODS

Data were from the 2023 and 2024 waves of the International Cannabis Policy Study (ICPS), which consists of repeat cross-sectional surveys conducted annually in Canada, United States, Australia, New Zealand, UK and Germany. The current study reported data from the UK sample only. Data were collected via self-completed web-based surveys in September-November in 2023 and 2024. Non-probability samples of respondents aged 16-65 were recruited through the Nielsen Consumer Insights Global Panel and their partners’ panels. Nielsen draws stratified random samples from the online panels, with quotas based on sex and age. For the UK sample, people who had used cannabis in the past 12 months were oversampled to ensure sufficient power for analyses. Upon completion, respondents received remuneration in accordance with their panel’s usual incentive structure. The American Association for Public Opinion Research cooperation rate (29), which is the percentage of respondents who completed the survey among all eligible respondents who accessed the survey link, was 55.1% in 2023 and 39.1% in 2024. The median survey time was 22 minutes.

The study was reviewed by and received ethics clearance through the University of Waterloo (ORE#31330) and the University of Bath (0513–586). Informed consent was obtained from all respondents and/or their legal guardian(s). A full description of the study methods can be found in the ICPS Technical Reports and methodology paper (30–32). The analysis plan for this paper was pre-registered on the Open Science Framework prior to data analysis (https://osf.io/j7ax6).

### Measures

#### Socio-demographic measures

Sex-at-birth, age, ethnicity/race, highest education level, perceived income adequacy, and region (Table 1). For “perceived income adequacy” and “ethnicity/race” those who answered, “Don’t know” or “Refuse to answer” were categorised to “Unstated”. Regions within England but not London were combined.

**Table 1:**
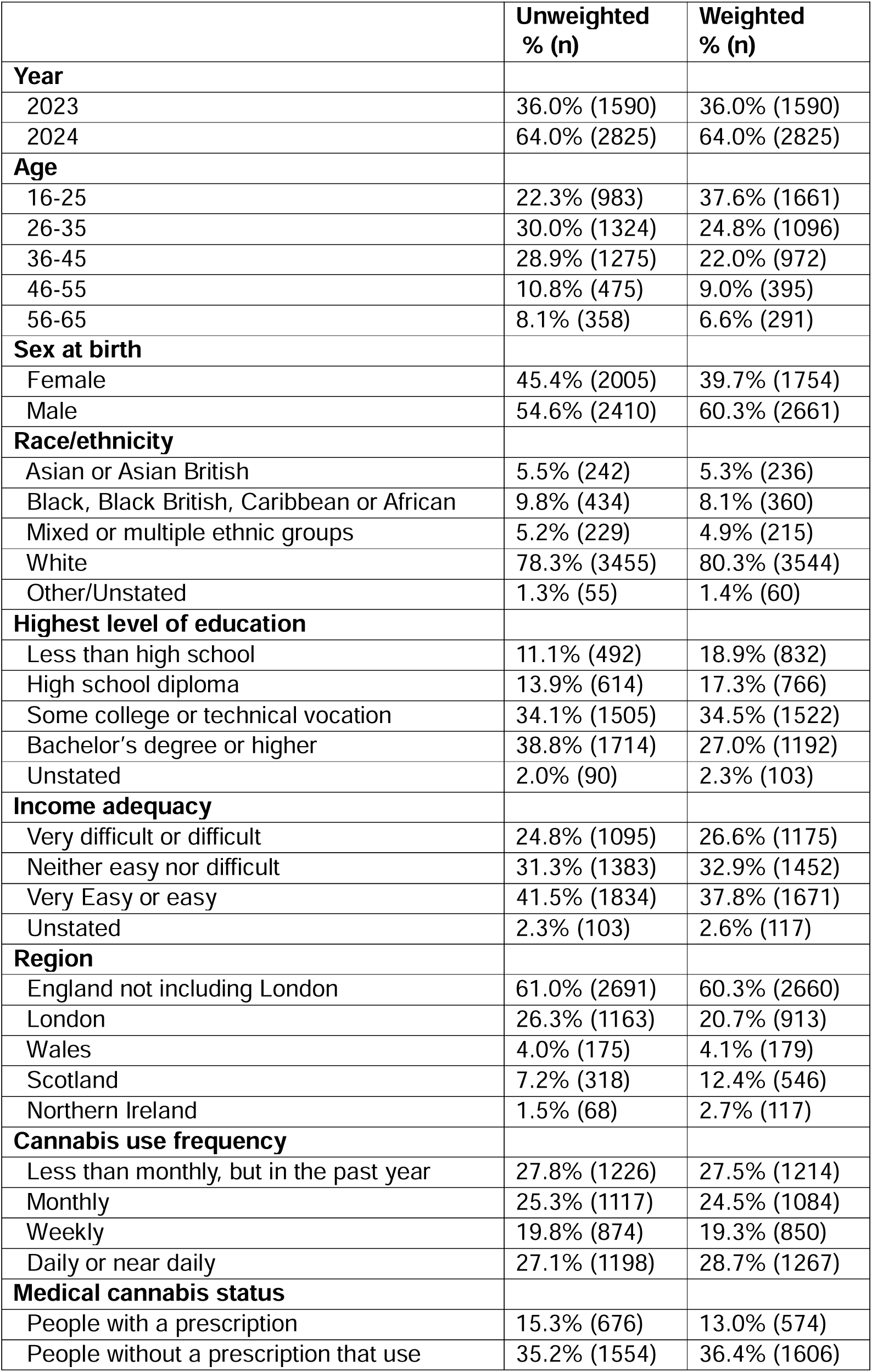
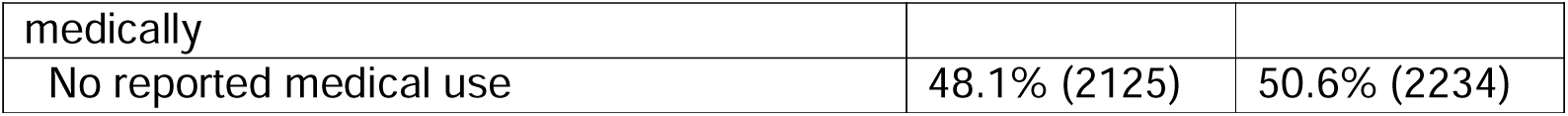
Unweighted and weighted sample characteristics among UK respondents that reported consuming cannabis in the past year in 2023-2024 (n=4,415).

#### High-risk use using the CUDIT-SF

High-risk use was assessed using the Cannabis Use Disorder Identification Test Revised Short Form (CUDIT-SF) (33). The CUDIT-SF is a three-item form that determines a positive screen to be two and above, which was found to correctly identify 78% of people with DSM-5 cannabis use disorder (33).

#### Medical cannabis use status

Medical cannabis use status was created from two questions. First, “Do you use cannabis for medical reasons, recreational reasons or both? By medical cannabis user, we mean someone who uses cannabis only to manage a medical condition.” (1=Medical use only, 2=Recreational use only, 3=Both recreational and medical use, 4=Don’t know). Second, “Did you receive a prescription to use medical cannabis at any time in the past 12 months?” (5=Yes, 6=No, 7=Don’t know). Responses to these questions were combined to create three exclusive categories: “People with a medical cannabis prescription” [5], “People without a medical cannabis prescription who use medically” [1,3,6,7], “No medical use” [2,4].

#### Asked for a medical cannabis prescription

Respondents were asked “Have you ever asked a licensed health professional for a prescription to use medical cannabis?” With response options “Yes” and “No” (No/Don’t know).

#### Cannabis product use and frequency

Respondents were asked whether they used each of nine product types in the past 12 months: dried flower, oils/liquids taken orally (drops or capsules), oil/liquid for vaping, edibles/foods, drinks, solid concentrates, hash/kief, tinctures, and topicals. Respondents who reported using a product were asked to provide their frequency of using each product. Drops and capsules were combined for past 12 months use but separated for frequency of use as per survey questioning. Responses were dichotomized to a “Monthly or more frequent use (MMF)” (Monthly/Weekly/Daily) versus “Other” (Less than monthly/Don’t know/Not applicable – i.e., those who did not use the product). Tinctures were not included in the analysis due to low cell counts and thus inability for models to converge.

#### Cannabis product sources

Respondents could select all that applied to the following question “In the past 12 months, have you gotten any type of cannabis from the following sources?” (I made or grew my own/From a family member or friend/From a dealer/Online/From a store or dispensary/Through medical prescription/Other). All were binary questions whereby selecting the source is “Yes” and the absence is “No”. “Other” was recategorised to existing options if provided in open text.

#### Source of medical cannabis prescription

Respondents who had reported receiving their cannabis through medical prescription were asked where they sourced it (NHS prescription/Private prescription/Patient registry).

#### Legally sourced medical cannabis

Respondents were asked “Overall, how much of the cannabis that you used in the past 12 months was from a legal medical prescription?” Respondents were categorised into four options “All” (100%), “Some” (1-99%), “None” (0%) and “Unstated” (I did not buy or pay for cannabis in the past 12 months/Don’t know/Refuse to answer).

All questions included “Don’t know” and “Refuse to answer” options and were excluded unless specified within the measures above.

### Analysis

The current analysis uses pooled UK data from 2023 (n=4,010) and 2024 (n=7,002). After removing respondents due to self-reported dishonesty determined by asking participants “Were you able to provide ‘honest’ answers about your cannabis use during the survey?” (n=256); poor data quality determined by those who did not select the current month (n=813); those who identified as intersex and an ‘other’/unstated sex due to insufficient cell counts for weighting (n=10); speeding (n=66); duplicate entries (n=272); and unstated region (n=22), 9,573 respondents were retained. The pooled analytic sample for the current study was 4,415 respondents, which includes data from people who reported using cannabis in the past 12 months (n_2023_=1,590; n_2024_=2,825). Missing data were removed using case-wise deletion for variables used in regression models: education (n=201; 2.1% of analytical sample), asking a licensed healthcare professional for a prescription (n=83; 1.9%) and medical cannabis status use (n=60; 1.4%).

Post-stratification sample weights were constructed to calibrate to population proportions as available in censuses and national benchmark surveys. Respondents were classified into age-by-sex-by-region groups, ethnicity-by-region groups, education groups, and age-by-sex-by-cannabis-use groups. Correspondingly grouped population proportion estimates were obtained from national government agencies (30, 32). A raking algorithm was applied to compute weights that are calibrated to these groupings. The SAS macro “RAKE_AND_TRIM_G4_V5” was used, with trimming to 5 (rescaled) if necessary. Estimates are weighted unless otherwise specified.

First, descriptive statistics were used to estimate the percentage of 1) people using cannabis for medical purposes with and without a prescription; 2) the use of cannabis products; and 3) the sources used to obtain cannabis – by medical cannabis use status. Second, Rao Scott chi-square tests of association were used between medical cannabis use status for source and product variables. Third, univariable and multivariable regression models were used to test the association between MMF product use and medical cannabis use status. Post-hoc sensitivity analyses that were not pre-registered were conducted where any MMF use (vs no MMF use) of all nine products was the outcome and where the proportion of cannabis used from a legal prescription was the predictor instead of receiving a prescription.

Three additional post-hoc analyses that were not pre-registered were conducted on 1) the percentage of perceived ease of accessing legal medical cannabis, including Rao Scott chi-square tests of association between medical cannabis use status; 2) multivariable regression models to test the association between asking for a medical cannabis prescription and 3) receiving a medical cannabis prescription, high-risk use and socio-demographic characteristics.

All models were adjusted for region, age, sex-at-birth, education, ethnicity/race, income adequacy, year and device used to complete survey. Adjusted risk ratios (aRR) are reported with 95% confidence intervals (95% CI). We used quasi-Poisson regression analyses with a log-link function to obtain adjusted risk ratios (34). Analyses were conducted using R version 4.4.1, with survey weighting and svyglm for models.

## RESULTS

Table 1 displays the sample characteristics of respondents who reported using cannabis in the past year. The sample were predominantly aged between 16 and 35 (62.4%), male-at-birth (60.3%), identified as White race/ethnicity (80.3%), and educated beyond high school (61.5%). Close to a quarter of people reported daily cannabis use (28.7%). A total of 13.0% reported receiving a medical cannabis prescription in the past year, 36.4% reported use of cannabis medically but without a medical cannabis prescription, and 50.6% did not report medical cannabis use.

### Medical cannabis sources

A total of 18.8% of people who used using cannabis reported ever asking for a medical cannabis prescription (Table 2). A total of 7.6% reported getting their cannabis from a medical prescription in the past year. When disaggregated by medical cannabis use status, 30.9% of people with a prescription, 7.1% of people without a prescription that used medically, and 2.2% of people that did not report medical use reported getting cannabis from a prescription (χ^2^=531.6, p<0.001). Small percentages of people who used cannabis reported to get their prescription from the NHS (2.8%) and privately (4.3%). Among people with a medical cannabis prescription, 2.8%, 63.3% and 10.7% reported that none, some and all of their cannabis was from a legal medical prescription, respectively. Greater percentages of people with a prescription screened positive for high-risk use (75.4%) compared to those without a prescription that used medically (45.9%), or those who reported no medical use (39.8%; χ^2^=233.9, p<0.001).

**Table 2:** Percentages of cannabis outcomes among people with and without a prescription to use medical cannabis and those who did not report medical use, 2023-2024 (n=4,415).

| | People who reported using cannabis in the past year (n=4,415) | People with a medical cannabis prescription in past year (n=676) | People without a prescription that use medically (n=1,554) | People who did not report medical use (n=2,125) | $\chi^2$, p-value |
| --- | --- | --- | --- | --- | --- |
|  | **Weighted % (unweighted n)** |  |  |  |  |
| **Ever asked for a medical cannabis prescription** |  |  |  |  |  |
| Yes | 18.8% (931) | 82.7% (570) | 13.6% (228) | 6.3% (132) | 1790.8, p<0.001 |
| **Sourced cannabis from medical prescription in past year** |  |  |  |  |  |
| Yes | 7.6% (370) | 30.9% (201) | 7.1% (112) | 2.2% (56) | 531.6, p<0.001 |
| **Source of medical prescription** |  |  |  |  |  |
| Through an NHS prescription | 2.8% (132) | 12.3% (78) | 2.6% (39) | 0.5% (15) | 558.3, p<0.001 |
| Through a private prescription | 4.3% (211) | 17.9% (114) | 3.8% (62) | 1.3% (24) |  |
| Through a patient registry | 0.4% (21) | 0.8% (9) | 0.4% (7) | 0.3% (5) |  |
| **How much of the cannabis used in the past year was from a legal medical prescription** |  |  |  |  |  |
| None (0%) | 35.2% (1492) | 2.8% (15) | 40.0% (585) | 40.9% (886) | 953.8, p<0.001 |
| Some (1-99%) | 21.9% (1169) | 63.3% (468) | 20.5% (385) | 12.4% (306) |  |
| All (100%) | 2.9% (144) | 10.7% (67) | 2.7% (39) | 1.0% (32) |  |
| Unstated | 40.0% (1610) | 23.3% (126) | 36.8% (545) | 45.7% (901) |  |
| **Sources of cannabis used in past year** |  |  |  |  |  |
| From a dealer (in person) | 53.4% (2321) | 48.6% (352) | 55.7% (836) | 53.6% (1114) | 8.6, p=0.07 |
| From a family member or friend | 43.1% (1960) | 32.3% (233) | 43.2% (683) | 46.4% (1026) | 36.9, p<0.05 |
| From a store or dispensary | 15.3% (792) | 35.9% (271) | 14.3% (243) | 10.9% (269) | 222.9, p<0.001 |
| Internet delivery service or mailed to me | 14.1% (713) | 28.8% (219) | 15.1% (261) | 9.8% (227) | 138.4, p<0.001 |
| I made or grew my own | 8.5% (443) | 28.8% (200) | 7.7% (135) | 4.0% (102) | 363.2, p<0.001 |
| **High-risk use from CUDIT-SF score** |  |  |  |  |  |
| Positive screen | 46.2% (2002) | 75.4% (497) | 45.9% (688) | 39.8% (807) | 233.9, p<0.001 |
| Negative screen | 53.8% (2413) | 24.6% (179) | 54.1% (866) | 60.3% (1318) |  |

In the past year, people who used cannabis reported getting cannabis from a dealer (53.4%), a family member/friend (43.1%), a physical store (15.3%), online (14.1%), and making or growing their own (8.5%). Greater percentages of people with a prescription reported making or growing their own cannabis (χ^2^=363.2, p<0.001), getting their cannabis online (χ^2^=138.4, p<0.001) and from a physical store (χ^2^=229.9, p<0.001). Greater percentages of people without a prescription – both those using medically and those not reporting medical use - reported getting their cannabis from a family member or friend (χ^2^=36.9, p<0.01). There was no difference between medical cannabis use status for getting cannabis from a dealer (χ^2^=8.6, p=0.07).

### Perceived access to medical cannabis

Two-thirds of people with a prescription reported perceiving access to a prescription as easy (65.7%), 17.7% as neither easy nor difficult, 15.1% as difficult, and 16.7% did not know (Supplemental Table 1). Greater percentages of people without a medical cannabis prescription but used medically and those who do not report medical use reported perceiving access difficult than people with a medical prescription (43.9% and 40.2% versus 15.1%, respectively; χ^2^=591.1, p<0.001).

### Product use

People with a medical cannabis prescription had greater percentages of past year use than people without a prescription but reporting medical use, and people who did not report medical use, for all products except dried flower (Figure 1). For dried flower, people with a prescription had lower percentages of past year use (all p<0.05). People with a medical cannabis prescription had greater percentages of MMF use in the past year than people without a prescription for medical cannabis but used medically for all products (all p<0.05) except dried flower (p>0.05) (Figure 2).

**Figure 1:**
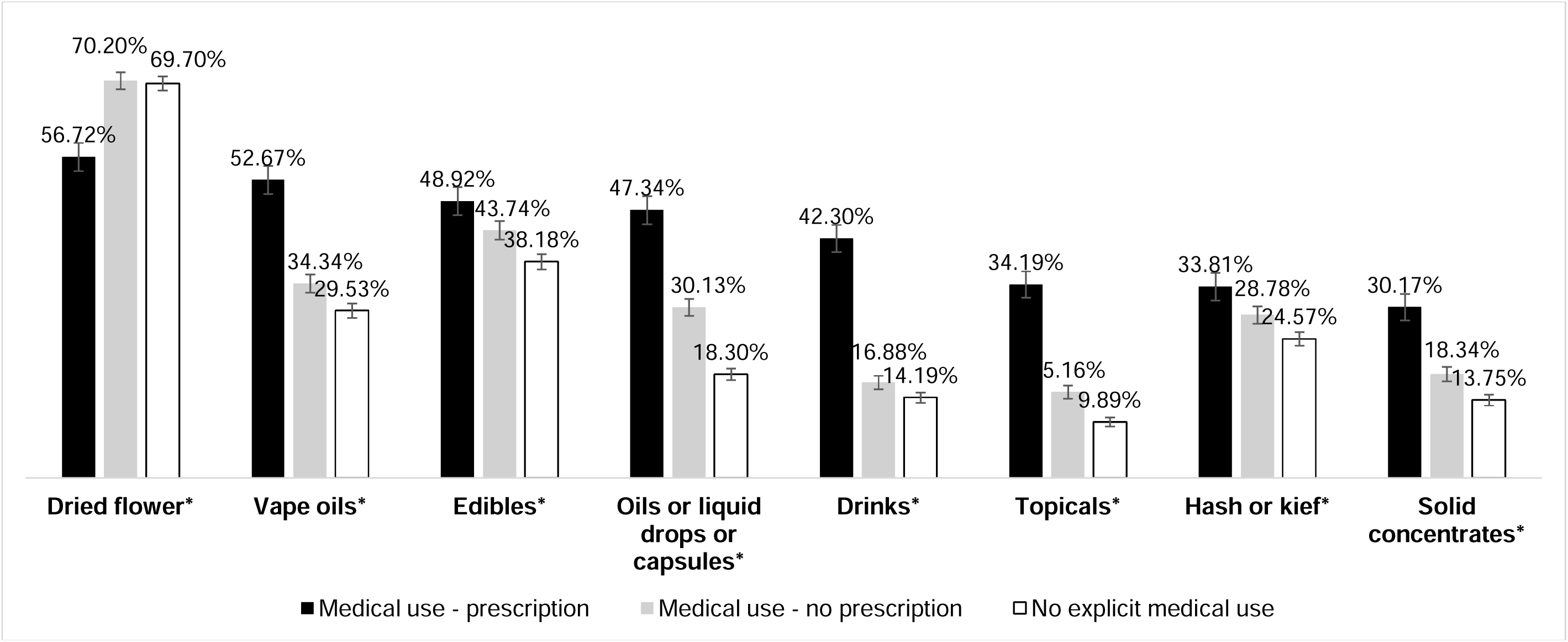
**Any use of each product in the past year among respondents with a medical cannabis prescription, those without a prescription but using medically, and those without reporting any medical use (n=4,415).** Footnote: * indicates a statistically significant difference between groups at the 5% level using a univariable quasi-Poisson regression analysis with a log-link function. Dried flower[a,b]; Vape oils[a,b,c]; Edibles[a,c]; Oils or liquid drops or capsules[a,b,c]; Drinks[a,b,c]; Topicals[a,b,c]; Hash or kief[a,c]; Solid concentrates[a,b,c] where a is p<0.05 between ‘medical use – prescription’ and ‘No medical use’, b is p<0.05 between ‘medical use – prescription’ and ‘medical use – no prescription’ and c is p<0.05 between ‘Medical use – no prescription’ and ‘No medical use’.

**Figure 2:**
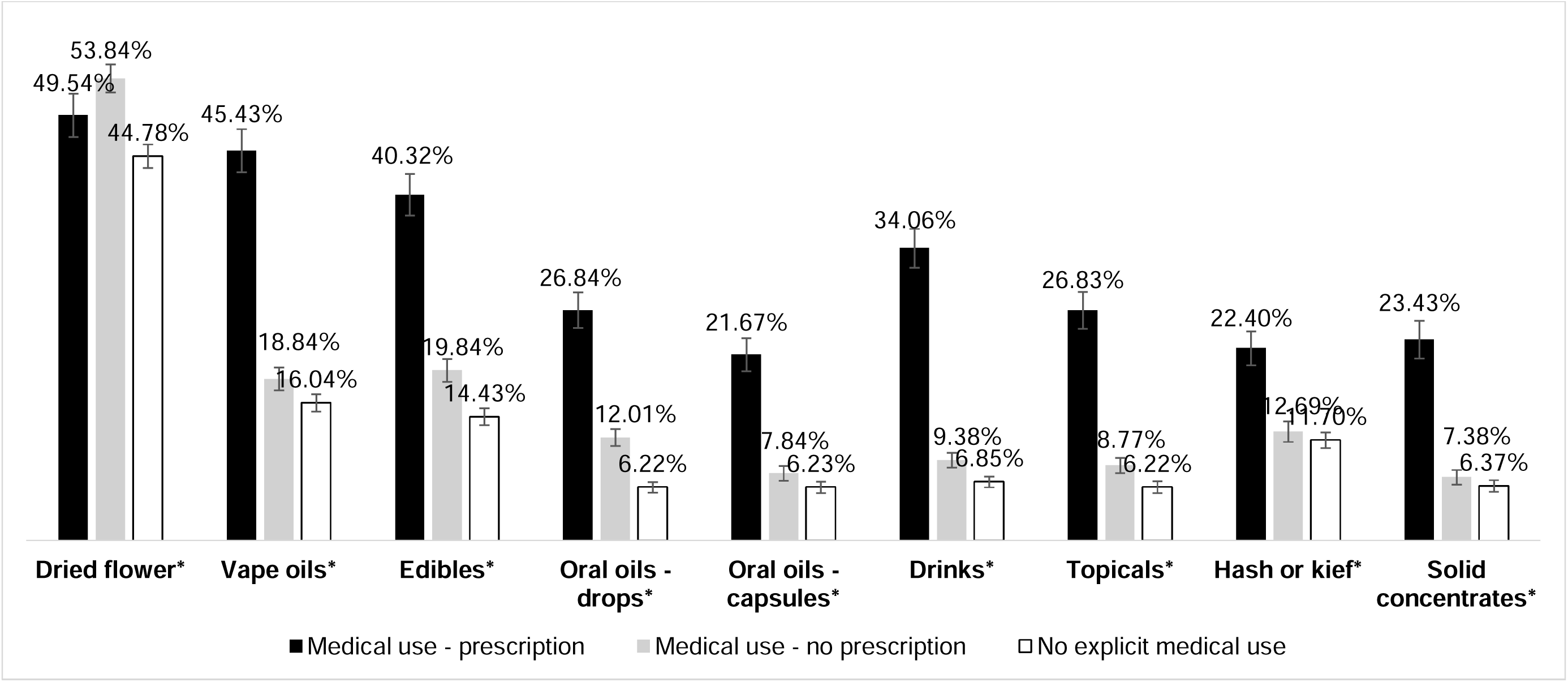
**Monthly or more frequent use of each product in the past year among respondents with a medical cannabis prescription, those without a prescription but using medically, and those without reporting any medical use (n=4,415).** Footnote: * indicates a statistically significant difference between groups at the 5% level using a univariable quasi-Poisson regression analysis with a log-link function. Dried flower[c]; Vape oils[a,b]; Edibles[a,b,c]; Oils or liquid drops[a,b,c], oils or liquid capsules[a,b]; Drinks[a,b,c]; Topicals[a,b,c]; Hash or kief[a,b]; Solid concentrates[a,b] where a is p<0.05 between ‘medical use – prescription’ and ‘No medical use’, b is p<0.05 between ‘medical use – prescription’ and ‘medical use – no prescription’ and c is p<0.05 between ‘Medical use – no prescription’ and ‘No medical use’.

Table 3 and Table 4 display regression analyses for MMF use. People with a medical cannabis prescription had a higher probability of MMF use of oils or liquid drops (aRR=3.51; 95% CI: 2.73-4.52), oil or liquid capsules (aRR=2.63; 1.99-3.47), vape oils (aRR=2.40; 2.03-2.84), edibles (aRR=2.41; 2.01-2.90), cannabis drinks (aRR=3.39; 2.69-4.26), solid concentrates (aRR=3.10; 2.29-4.21), hash or kief (aRR=1.82; 1.44-2.30), and topicals (aRR=3.42; 2.58-4.51) than people not reporting medical use, after adjusting for covariates. People without a prescription who reported medical use had a higher probability of MMF use of dried flower (aRR=1.18; 1.09-1.29), oils or liquid drops (aRR=1.93; 1.50-2.49), vape oils (aRR=1.27; 1.06-1.52), edibles (aRR=1.49; 1.24-1.79), cannabis drinks (aRR=1.44; 1.11-1.87) and topicals (aRR=1.46; 1.09-1.97) than people not reporting medical use, after adjusting for covariates. Supplemental Tables 2 and 3 display results from the sensitivity analyses, which demonstrated a similar pattern of results.

**Table 3:** Weighted regression analyses on the monthly or more frequent use of cannabis products and medical cannabis prescriptions (n=4,273).

|  | Monthly or more frequent use of... (vs Other) |  |  |  |  |
| --- | --- | --- | --- | --- | --- |
|  | Dried flower | Oil or liquid drops | Oil or liquid capsules | Vape oils | Edibles |
|  | aRR (95% CI) |  |  |  |  |
| **Medical cannabis use status** (vs *No medical use*) |  |  |  |  |  |
| People with a prescription | 1.09 (0.97, 1.23) | **3.51 (2.73, 4.52)** | **2.63 (1.99, 3.47)** | **2.40 (2.03, 2.84)** | **2.41 (2.01, 2.90)** |
| People without a prescription that use medically | **1.18 (1.09, 1.29)** | **1.93 (1.50, 2.49)** | 1.29 (0.96, 1.74) | **1.27 (1.06, 1.52)** | **1.49 (1.24, 1.79)** |
| **Age** (vs 56-65) |  |  |  |  |  |
| 16-25 | 0.93 (0.80, 1.09) | 0.87 (0.58, 1.33) | **2.59 (1.30, 5.16)** | **2.70 (1.74, 4.20)** | **2.29 (1.56, 3.37)** |
| 26-35 | 1.13 (0.99, 1.31) | 1.12 (0.77, 1.63) | **3.03 (1.59, 5.78)** | **2.48 (1.61, 3.82)** | **2.03 (1.39, 2.97)** |
| 36-45 | **1.16 (1.01, 1.33)** | 1.38 (0.97, 1.97) | **3.11 (1.64, 5.89)** | **2.39 (1.55, 3.67)** | **2.08 (1.43, 3.03)** |
| 46-55 | 1.07 (0.91, 1.26) | 0.82 (0.53, 1.27) | **2.77 (1.37, 5.61)** | 1.23 (0.75, 2.03) | 1.08 (0.69, 1.70) |
| **Sex at birth** (vs *Male*) |  |  |  |  |  |
| Female | **0.82 (0.76, 0.89)** | 1.08 (0.89, 1.32) | **0.79 (0.62, 0.99)** | **0.74 (0.63, 0.85)** | 0.87 (0.74, 1.01) |
| **Race/ethnicity** (vs *White*) |  |  |  |  |  |
| Asian or Asian British | **0.68 (0.54, 0.86)** | 0.99 (0.63, 1.59) | 1.32 (0.80, 2.16) | **1.36 (1.05, 1.76)** | 0.86 (0.60, 1.22) |
| Black, Black British, Caribbean or African | 1.10 (0.97, 1.25) | 1.09 (0.77, 1.54) | 1.08 (0.77, 1.52) | 0.93 (0.75, 1.15) | 0.93 (0.75, 1.16) |
| Mixed or multiple ethnic groups | 1.13 (0.95, 1.34) | 0.99 (0.63, 1.59) | 0.95 (0.56, 1.61) | 0.84 (0.63, 1.12) | 0.97 (0.72, 1.31) |
| Other/Unstated | 0.89 (0.55, 1.39) | **0.13 (0.02, 0.92)** | **0.11 (0.02, 0.83)** | 1.47 (0.84, 2.57) | 1.17 (0.58, 2.36) |
| **Highest level of education** (vs *Less than high school*) |  |  |  |  |  |
| High school diploma | 0.94 (0.82, 1.07) | **2.03 (1.23, 3.34)** | 1.04 (0.56, 1.92) | **1.54 (1.08, 2.19)** | 1.38 (0.97, 1.96) |
| Some college or technical vocation | **0.80 (0.71, 0.90)** | **1.91 (1.20, 3.02)** | 1.30 (0.76, 2.21) | **1.47 (1.07, 2.00)** | 1.26 (0.92, 1.73) |
| Bachelor's degree or higher | **0.82 (0.73, 0.92)** | **2.06 (1.30, 3.26)** | 1.40 (0.84, 2.34) | **1.50 (1.10, 2.05)** | **1.42 (1.04, 1.93)** |
| **Income adequacy** (vs *Very/Difficult*) |  |  |  |  |  |
| Neither easy nor difficult | 0.99 (0.90, 1.09) | 0.88 (0.65, 1.19) | 0.90 (0.63, 1.29) | 1.21 (0.97, 1.51) | 1.23 (0.97, 1.56) |
| Very Easy or easy | 0.96 (0.87, 1.07) | 1.12 (0.85, 1.46) | **1.39 (1.00, 1.93)** | **1.34 (1.10, 1.64)** | **1.62 (1.31, 2.00)** |
| Unstated | 1.07 (0.81, 1.41) | 0.24 (0.06, 1.00) | 0.13 (0.02, 1.00) | 0.95 (0.39, 2.29) | 1.25 (0.65, 2.40) |
| **Region** (*vs England no London*) |  |  |  |  |  |
| London | 1.05 (0.96, 1.15) | 0.99 (0.81, 1.21) | **1.33 (1.06, 1.67)** | **1.20 (1.04, 1.38)** | 1.01 (0.87, 1.18) |
| Wales | 1.14 (0.95, 1.36) | 1.07 (0.64, 1.79) | 1.34 (0.73, 2.45) | 0.80 (0.45, 1.44) | **0.55 (0.35, 0.87)** |
| Scotland | **1.16 (1.02, 1.32)** | 0.72 (0.48, 1.08) | 1.19 (0.76, 1.85) | 1.01 (0.76, 1.34) | 1.02 (0.78, 1.34) |
| Northern Ireland | 0.97 (0.73, 1.29) | 0.87 (0.43, 1.79) | 1.13 (0.51, 2.51) | 1.55 (0.94, 2.55) | 1.07 (0.64, 1.80) |
| **Year** (*vs 2023*) |  |  |  |  |  |
| 2024 | 1.01 (0.93, 1.10) | 1.05 (0.84, 1.30) | 1.10 (0.82, 1.46) | 1.08 (0.92, 1.27) | 0.95 (0.80, 1.13) |
| **Survey device used** (*vs Smartphone*) |  |  |  |  |  |
| Tablet | 0.85 (0.66, 1.09) | 1.03 (0.58, 1.84) | 1.24 (0.63, 2.41) | 1.33 (0.86, 2.06) | 0.61 (0.32, 1.13) |
| Computer | 0.92 (0.84, 1.01) | 1.19 (0.96, 1.48) | 1.32 (0.99, 1.74) | 1.04 (0.89, 1.22) | 0.91 (0.77, 1.09) |

**Table 4:** Weighted regression analyses on the monthly or more frequent use of cannabis products and medical cannabis prescriptions (n=4,273).

|  | Monthly or more frequent use of... (vs Other) |  |  |  |
| --- | --- | --- | --- | --- |
|  | Drinks | Solid Concentrates | Hash or kief | Topicals |
|  | aRR (95% CI) |  |  |  |
| **Medical cannabis use status** (vs *No medical use*) |  |  |  |  |
| People with a prescription | **3.39 (2.69, 4.26)** | **3.10 (2.29, 4.21)** | **1.82 (1.44, 2.30)** | **3.42 (2.58, 4.51)** |
| People without a prescription that use medically | **1.44 (1.11, 1.87)** | 1.19 (0.87, 1.63) | 1.09 (0.86, 1.39) | **1.46 (1.09, 1.97)** |
| **Age** (vs 56-65) |  |  |  |  |
| 16-25 | **3.88 (1.48, 10.19)** | **5.87 (2.19, 15.77)** | **1.70 (1.07, 2.70)** | **2.70 (1.43, 5.09)** |
| 26-35 | **4.83 (1.88, 12.44)** | **5.63 (2.15, 14.81)** | 1.46 (0.92, 2.34) | **2.46 (1.33, 4.58)** |
| 36-45 | **4.74 (1.86, 12.05)** | **6.81 (2.61, 17.78)** | **2.16 (1.39, 3.34)** | **2.96 (1.62, 5.40)** |
| 46-55 | **2.86 (1.06, 7.69)** | **2.92 (1.04, 8.17)** | 1.53 (0.92, 2.53) | 1.48 (0.74, 2.94) |
| **Sex at birth** (vs *Male*) |  |  |  |  |
| Female | **0.74 (0.61, 0.90)** | **0.77 (0.60, 0.99)** | **0.62 (0.50, 0.77)** | 0.98 (0.79, 1.23) |
| **Race/ethnicity** (vs *White*) |  |  |  |  |
| Asian or Asian British | 1.15 (0.76, 1.75) | 0.90 (0.54, 1.51) | 1.12 (0.75, 1.66) | 1.44 (0.92, 2.25) |
| Black, Black British, Caribbean or African | 1.11 (0.86, 1.44) | 0.97 (0.65, 1.46) | 1.09 (0.76, 1.57) | 1.12 (0.79, 1.59) |
| Mixed or multiple ethnic groups | 0.86 (0.60, 1.25) | 0.99 (0.60, 1.63) | 1.21 (0.82, 1.78) | 0.73 (0.48, 1.12) |
| Other/Unstated | 0.48 (0.17, 1.35) | 0.27 (0.06, 1.15) | 1.85 (0.90, 3.79) | 0.57 (0.16, 2.00) |
| **Highest level of education** (vs *Less than high school*) |  |  |  |  |
| High school diploma | 1.70 (0.88, 3.30) | 1.09 (0.62, 1.92) | 1.33 (0.90, 1.96) | 1.65 (0.87, 3.11) |
| Some college or technical vocation | **2.12 (1.19, 3.75)** | 1.00 (0.61, 1.67) | 1.25 (0.88, 1.78) | 1.59 (0.89, 2.83) |
| Bachelor's degree or higher | **3.15 (1.79, 5.54)** | 1.00 (0.62, 1.61) | 0.90 (0.63, 1.29) | **1.85 (1.06, 3.24)** |
| **Income adequacy** (vs *Very/Difficult*) |  |  |  |  |
| Neither easy nor difficult | 0.99 (0.72, 1.38) | 1.03 (0.70, 1.52) | 0.83 (0.63, 1.09) | 1.16 (0.81, 1.67) |
| Very Easy or easy | **1.46 (1.09, 1.97)** | 1.18 (0.86, 1.62) | 0.88 (0.68, 1.12) | **1.43 (1.03, 1.98)** |
| Unstated | 0.70 (0.23, 2.15) | 1.33 (0.46, 3.79) | 0.47 (0.15, 1.42) | 0.98 (0.24, 4.00) |
| **Region** (*vs England no London*) |  |  |  |  |
| London | **1.43 (1.19, 1.73)** | **1.32 (1.05, 1.67)** | 1.10 (0.89, 1.36) | 1.20 (0.96, 1.50) |
| Wales | 1.37 (0.81, 2.33) | 0.63 (0.34, 1.18) | **0.40 (0.21, 0.74)** | 1.13 (0.61, 2.07) |
| Scotland | 0.75 (0.47, 1.20) | 1.04 (0.62, 1.74) | 1.13 (0.79, 1.63) | 0.91 (0.58, 1.43) |
| Northern Ireland | 1.12 (0.54, 2.32) | 1.27 (0.42, 3.81) | 0.94 (0.43, 2.06) | 2.01 (0.95, 4.25) |
| **Year** (*vs 2023*) |  |  |  |  |
| 2024 | 0.99 (0.80, 1.23) | 0.93 (0.70, 1.24) | 0.94 (0.75, 1.17) | 0.81 (0.63, 1.06) |
| **Survey device used** (*vs Smartphone*) |  |  |  |  |
| Tablet | 1.14 (0.57, 2.28) | 0.78 (0.26, 2.33) | 0.65 (0.31, 1.35) | 1.06 (0.49, 2.29) |
| Computer | 0.96 (0.78, 1.17) | 1.19 (0.92, 1.54) | 1.01 (0.81, 1.25) | **1.37 (1.06, 1.76)** |

### Characteristics of those who asked for and received a medical cannabis prescription

People who ever *asked* for a medical prescription and people who *received* a medical prescription in the past year had a higher probability of screening positive for high-risk use. (Supplemental Table 4). People who had a higher probability of ever asking for a medical cannabis prescription and receiving a medical cannabis prescription in the past year were those aged 26-45, male-at-birth, educated to some college/technical vocation or higher, and residing in London.

## DISCUSSION

To our knowledge, this is the first study to examine the products and sources used among people with and without a medical cannabis prescription in the UK. The study has four primary findings: First, close to one in two people who used cannabis reported using cannabis for medical purposes, but only one in eight reported receiving a medical cannabis prescription in the past year. Second, there is a diversity of sources used to obtain cannabis; only one in 10 people with a prescription obtained all their cannabis from a legal medical prescription and a dealer was the most used source. Third, people who reported receiving a medical cannabis prescription had a higher probability of consuming processed non-flower products monthly or more frequently than those without a prescription or those who did not report medical use. Fourth, people who ever asked for a medical prescription and people with a medical prescription in the past year had a higher probability of screening positive for high-risk use.

Among those who reported using cannabis for medical purposes – a larger proportion reported using medically without a prescription than those who reported receiving a prescription in the past year. A similar disparity was demonstrated in a report by the Centre for Medicinal Cannabis, which found that 1.4 million people in the UK were consuming cannabis for medical purposes yet during the same period of the survey, fewer than 100 private prescriptions had been issued (35). Another survey in 2022 concluded a higher figure - that 1.7 million people were consuming illegal cannabis to treat a health condition (11). While our study suggests a larger proportion of people using cannabis for reported medical purposes are accessing legal medical cannabis, there still appears to be an unmet need of those self-medicating without access to formal medical guidance or oversight. However, it may be the case that some people using for reported medical use are unlikely to be approved for a prescription if they do not have an adequate need or if all other treatments have not been exhausted, as per UK guidance. Furthermore, we found that the number reporting to access cannabis via NHS prescription was higher than expected, suggesting that some people accessing cannabis via prescription may misunderstand this distinction between public and private prescriptions.

There is a diversity of sources used to obtain cannabis, even among those that had received a prescription. Indeed, only one in 10 people that had a medical cannabis prescription sourced all of their cannabis from a legal prescription. Most only obtained *some* of their cannabis from a prescription and the most common source used by those with a prescription was a dealer. It may be that those who have a prescription also consume cannabis for non-medical purposes (i.e., recreationally) and use a different source for each purpose.

Alternatively, they may experience problems with legal medical cannabis that do not meet consumer needs (e.g., based on availability, product, and/or price). In a qualitative study exploring the experiences of UK medical cannabis patients, patients reported prohibitive costs and supply issues, noting that prescriptions went out of stock or products were low quality (e.g., mould) (16). Moreover, in the current study, close to half of people without a prescription that reported medical use reported perceiving access to a prescription to be difficult. This is consistent with a survey exploring why UK respondents who obtained their medical cannabis illegally did so rather than sourcing legally; the most common reason cited was the presumed difficulties in access (11). Our study demonstrates that there is a “mixed economy” in terms of sourcing cannabis, whether that be for medical or recreational purposes. Further research should explore the reasons why people who use cannabis medically source from outside the legal market and what the illegal market provides that the legal medical market does not.

People with a medical cannabis prescription had a higher probability of frequently consuming processed products, which typically contain higher potencies than dried flower, than those who do not report medical use. Only certain types of products are permitted in the legal medical market and so while one may expect to see more ‘medical’ products being consumed by those with a prescription (e.g., oils and capsules), those with a prescription are also frequently consuming products that are not permitted on the legal medical market (e.g., hash or solid concentrates). It is important to note that people with prescriptions may also be consuming cannabis for recreational purposes, of which the other products may have been obtained illegally. Regardless, evidence points to an association between higher potency cannabis (and frequent use) with increased risk of cannabis use disorder (20, 21, 36). As estimates suggest cannabis use disorder affects one in four people who use medical cannabis (37), this is an important health outcome that should be discussed among people accessing medical cannabis. Indeed, in the current study, respondents with a prescription had a higher probability of screening positive for high-risk use. Contact with healthcare professionals when accessing prescriptions for medical cannabis could offer an opportunity to discuss risks of cannabis use disorder, and to consider how the type of product used, frequency of use and total THC exposure might influence risk of cannabis use disorder and other health outcomes. For example, safer use limits on THC consumption could be used to minimise the risk of cannabis use disorder (38).

## Limitations

This study is subject to limitations common to survey research. Respondents were recruited using non–probability-based sampling and are therefore not necessarily nationally representative; however, the data were weighted by age, sex, region, ethnicity, education and cannabis use to align with national distributions.

Self-reported data are subject to social desirability biases. Cannabis is an illegal drug in the UK; therefore, patterns of cannabis use may be underreported or misrepresented. Indeed, while medical cannabis is legal in the UK, there is stigma surrounding use of the drug for medical purposes (39). However, the survey included a data integrity question wherein those who reported not answering questions honestly were excluded. In addition, this survey was self-administered online, which compared to interviewer assisted surveys, can reduce social desirability biases by providing anonymity (40). Finally, cannabis use for medical purposes was self-reported and may not reflect medical use or need as identified by a healthcare professional.

## Conclusion

More people are using cannabis for medical purposes without a prescription than those with one. It appears that medical cannabis prescriptions in the UK may not be addressing the needs of the people reporting consuming cannabis medically, both for those with and without a prescription, as the illegal market continues to be utilised. Further, people with a prescription are using processed products more frequently than those without a prescription, which typically contain higher potencies than dried flower. Contact with healthcare professionals when accessing prescriptions for medical cannabis use should discuss risks of related to higher potency products, frequency of use and total THC exposure might influence risk of cannabis use disorder and other health outcomes.

## Author contributions

EW: Conceptualization (equal); formal analysis (lead); writing – original draft (lead); writing – review and editing (equal). TPF: Conceptualization (equal); formal analysis (support); writing – original draft (support); writing – review and editing (equal). DH: Conceptualization (equal); data curation (lead); writing – review and editing (equal). All authors contributed to and have approved the final manuscript.

## Data Availability

All data produced in the present study are available upon reasonable request to the authors.

https://cannabisproject.ca/methods

## Acknowledgements

EW is the recipient of fellowship funding from the UK Society for the Study of Addiction (SSA). TF is funded by a UKRI Future Leaders Fellowship [MR/Y017560/1]. The ICPS was supported by a Canadian Institutes of Health Research Project Bridge Grant [PJT-153342] (DH), a Canadian Institutes of Health Research Project Grant [PJT-153342] (DH), and UKRI [MR/Y017560/1] (TF).

## Declaration of interests

The authors have no conflicts of interest to declare. DH has served as a paid Expert Witness on behalf of public health authorities in response to industry legal challenges to cannabis regulations in Canada.

## Ethics statement

The study was reviewed by and received ethics clearance through the University of Waterloo (ORE#31330) and the University of Bath (0513–586).

## Data availability statement

The data that support the findings of this study are available from the International Cannabis Policy Study (www.cannabisproject.ca) but restrictions apply to the availability of these data and so are not publicly available. Data are however available from the authors upon reasonable request.

## Supplemental Files

**Supplemental Table 1: Percentages of perceived ease of access to get a medical cannabis prescription among people who use cannabis with and without receiving a medical prescription.**

**Supplemental Table 2: Weighted regression analyses on the monthly or more frequent use of at least one of the nine cannabis products assessed (n=4,273).**

**Supplemental Table 3: Univariable and multivariable regression analyses on the monthly or more frequent use of nine cannabis products and the use of cannabis from legal medical prescription (n=4,206).**

**Supplemental Table 4: Multivariable regression analyses on the characteristics of people who have ever asked for a medical cannabis prescription and who have ever received a medical prescription for cannabis in the past year (n=4,325).**

## Notes

### Author Declarations

The study was reviewed by and received ethics clearance through the University of Waterloo (ORE#31330) and the University of Bath (0513-586). Informed consent was obtained from all respondents and/or their legal guardian(s).

## References

1. Office for National Statistics. Drug misuse in England and Wales: year ending March 2024 2024 [Available from: https://www.ons.gov.uk/peoplepopulationandcommunity/crimeandjustice/articles/drugmisuseinenglandandwales/yearendingmarch2024.

2. National Institute for Health and Care Excellence. Cannabis-based medicinal products 2019 [updated 22 Mar 2021. Available from: https://www.nice.org.uk/guidance/NG144.

3. Freeman TP, Morgan C, Hindocha C. Strengthening the evidence for medicinal cannabis and cannabinoids. British Medical Journal Publishing Group; 2019.

4. Lake S, Walsh Z, Kerr T, Cooper ZD, Buxton J, Wood E, et al. Frequency of cannabis and illicit opioid use among people who use drugs and report chronic pain: a longitudinal analysis. PLoS medicine. 2019;16(11):e1002967.

5. Lucas P, Walsh Z. Medical cannabis access, use, and substitution for prescription opioids and other substances: a survey of authorized medical cannabis patients. International Journal of Drug Policy. 2017;42:30–5.

6. Walsh Z, Callaway R, Belle-Isle L, Capler R, Kay R, Lucas P, et al. Cannabis for therapeutic purposes: patient characteristics, access, and reasons for use. International Journal of Drug Policy. 2013;24(6):511–6.

7. Walsh Z, Gonzalez R, Crosby K, Thiessen MS, Carroll C, Bonn-Miller MO. Medical cannabis and mental health: A guided systematic review. Clinical psychology review. 2017;51:15–29.

8. Kosiba JD, Maisto SA, Ditre JW. Patient-reported use of medical cannabis for pain, anxiety, and depression symptoms: Systematic review and meta-analysis. Social science & medicine. 2019;233:181–92.

9. Sciences NAo, Medicine, Division M, Health BoP, Practice PH, Marijuana CotHEo, et al. The health effects of cannabis and cannabinoids: the current state of evidence and recommendations for research. 2017.

10. Couch D, Couch D, Chb M, Mba M. Left behind: the scale of illegal cannabis use for medicinal intent in the UK. Centre Med Cannabis. 2020.

11. Erridge S, Troup L, Sodergren MH. Illicit cannabis use to self-treat chronic health conditions in the United Kingdom: cross-sectional study. JMIR Public Health and Surveillance. 2024;10:e57595.

12. Arjun M, Verma V, Bhoopathi S, Mishra A, Venkatesh M. Medical cannabis: Regulatory review in United States, European Union and United Kingdom. Journal of Forensic and Legal Medicine. 2025:102947.

13. UK Parliament. Cannabis: Medical Treatments 2024 [Available from: https://questions-statements.parliament.uk/written-questions/detail/2024-12-04/17970.

14. Case P. The NICE guideline on medicinal cannabis: keeping Pandora’s box shut tight? Medical Law Review. 2020;28(2):401–11.

15. Heeg H, Morari A, Lynskey M, Turner JJ. An exploration of medical professionals’ attitudes, perceived knowledge and concerns around medical cannabis in the United Kingdom. Drug Science, Policy and Law. 2024;10:20503245241283699.

16. Wilson HB, McGrath LM. “It’s a big added stress on top of being so ill”: The challenges facing people prescribed cannabis in the UK. International Journal of Drug Policy. 2023;122:104220.

17. Pertwee RG. Handbook of cannabis: Oxford University Press, USA; 2014.

18. Freeman TP, Craft S, Wilson J, Stylianou S, ElSohly M, Di Forti M, et al. Changes in delta_-_9_-_tetrahydrocannabinol (THC) and cannabidiol (CBD) concentrations in cannabis over time: systematic review and meta_-_analysis. Addiction. 2021;116(5):1000–10.

19. Di Forti M, Quattrone D, Freeman TP, Tripoli G, Gayer-Anderson C, Quigley H, et al. The contribution of cannabis use to variation in the incidence of psychotic disorder across Europe (EU-GEI): a multicentre case-control study. The Lancet Psychiatry. 2019;6(5):427–36.

20. Freeman T, Winstock A. Examining the profile of high-potency cannabis and its association with severity of cannabis dependence. Psychological medicine. 2015;45(15):3181–9.

21. Petrilli K, Ofori S, Hines L, Taylor G, Adams S, Freeman TP. Association of cannabis potency with mental ill health and addiction: a systematic review. The Lancet Psychiatry. 2022;9(9):736–50.

22. Di Forti M, Marconi A, Carra E, Fraietta S, Trotta A, Bonomo M, et al. Proportion of patients in south London with first-episode psychosis attributable to use of high potency cannabis: a case-control study. The Lancet Psychiatry. 2015;2(3):233–8.

23. Hall W, Degenhardt L. High potency cannabis: a risk factor for dependence, poor psychosocial outcomes, and psychosis. BMJ 350, h1205. 2015.

24. Hindocha C, Freeman TP, Ferris JA, Lynskey MT, Winstock AR. No smoke without tobacco: a global overview of cannabis and tobacco routes of administration and their association with intention to quit. Frontiers in psychiatry. 2016;7:104.

25. Hammond D, Wadsworth E, Reid JL, Burkhalter R. Prevalence and modes of cannabis use among youth in Canada, England, and the US, 2017 to 2019. Drug and alcohol dependence. 2021;219:108505.

26. Medicines and Healthcare Products Regulatory Agency. Supply unlicensed medicinal products (specials) 2014 [updated 29 Jan 2025. Available from: https://www.gov.uk/government/publications/supply-unlicensed-medicinal-products-specials.

27. Advisory Council on the Misuse of Drugs. Cannabis-based products for medicinal use (CBPMs) in humans 2020 [Available from: https://assets.publishing.service.gov.uk/media/5fc0de5b8fa8f559ded502cb/OFFICIAL_Published_version_-_ACMD_CBPMs_report_27_November_2020_FINAL.pdf.

28. Advisory Council on the Misuse of Drugs. Cannabis-based products for medicinal use: call for evidence 2025 [Available from: https://www.gov.uk/government/calls-for-evidence/cannabis-based-products-for-medicinal-use/cannabis-based-products-for-medicinal-use-call-for-evidence.

29. American Association for Public Opinion Research. Standard definitions: Final dispositions of case codes and outcome rates for surveys 2016 [Available from: https://www.aapor.org/AAPOR_Main/media/publications/Standard-Definitions20169theditionfinal.pdf.

30. Fataar F, Iraniparast M, Rynard V, Burkhalter R, Hammond D. International Cannabis Policy Study Technical Report - Wave 7 (2024). University of Waterloo, Ontario, Canada: University of Waterloo; 2025.

31. Hammond D, Goodman S, Wadsworth E, Rynard V, Boudreau C, Hall W. Evaluating the impacts of cannabis legalization: the International Cannabis Policy Study. International Journal of Drug Policy. 2020;77:102698.

32. Iraniparast M, Rynard V, Burkhalter R, Corsetti D, Hammond D. International Cannabis Policy Study Technical Report - Wave 6 (2023). University of Waterloo, Ontario, Canada: University of Waterloo; 2023.

33. Bonn-Miller MO, Heinz AJ, Smith EV, Bruno R, Adamson S. Preliminary development of a brief cannabis use disorder screening tool: the cannabis use disorder identification test short-form. Cannabis and cannabinoid research. 2016;1(1):252–61.

34. Norton EC, Dowd BE, Maciejewski ML. Odds ratios—current best practice and use. Jama. 2018;320(1):84–5.

35. Nutt D, Bazire S, Phillips LD, Schlag AK. So near yet so far: why won’t the UK prescribe medical cannabis? BMJ open. 2020;10(9):e038687.

36. Craft S, Winstock A, Ferris J, Mackie C, Lynskey MT, Freeman TP. Characterising heterogeneity in the use of different cannabis products: latent class analysis with 55 000 people who use cannabis and associations with severity of cannabis dependence. Psychological medicine. 2020;50(14):2364–73.

37. Dawson D, Stjepanović D, Lorenzetti V, Cheung C, Hall W, Leung J. The prevalence of cannabis use disorders in people who use medicinal cannabis: A systematic review and meta-analysis. Drug and alcohol dependence. 2024;257:111263.

38. Thorne RL, Lawn W, Petrilli K, Trinci K, Borissova A, Ofori S, et al. Estimating thresholds for risk of cannabis use disorder using standard delta-9-tetrahydrocannabinol (THC) units. Addiction (Abingdon, England). 2025.

39. Troup LJ, Erridge S, Ciesluk B, Sodergren MH. Perceived stigma of patients undergoing treatment with cannabis-based medicinal products. International Journal of Environmental Research and Public Health. 2022;19(12):7499.

40. Hays RD, Liu H, Kapteyn A. Use of Internet panels to conduct surveys. Behavior research methods. 2015;47(3):685–90.

