## Supplemental Files 1-4 for "Medical cannabis in the UK: prescriptions, sources, products, and high-risk use"

**Supplemental Table 1: Percentages of perceived ease of access to get a medical cannabis prescription among people who use cannabis with and without receiving a medical prescription.**

|  | **People who reported using cannabis in the past year (n=4,415)** | **People with a medical cannabis prescription in past year (n=676)** | **People without a prescription that use medically (n=1,554)** | **People who did not report medical use (n=2,125)** | **χ^2^ , p-value** |
| --- | --- | --- | --- | --- | --- |
|  | **Weighted % (unweighted n)** | | | |  |
| **Perceived ease of access to get a medical cannabis prescription in the respondent’s city or town** |  |  |  |  | 591.1, p<0.05 |
| Very or fairly easy | 27.6% (1321) | 65.7% (446) | 24.7% (413) | 20.2% (457) |  |
| Neither easy nor difficult | 17.4% (731) | 17.7% (117) | 18.2% (276) | 16.6% (329) |  |
| Very or fairly difficult | 38.2% (1573) | 15.1% (98) | 43.9% (653) | 40.2% (814) |  |
| Don’t know | 16.7% (745) | 1.5% (11) | 13.1% (210) | 23.0% (515) |  |

Perceived ease of accessing legal medical cannabis: Respondents were asked “How easy or difficult would it be for you to get a medical cannabis prescription in the city or town where you live?” Response options were categorised to “Easy” (Very easy/Fairly easy), “Neither easy nor difficult” and “Difficult” (Very difficult/Fairly difficult), and “Don’t know”.

**Supplemental Table 2: Weighted regression analyses on the monthly or more frequent use of at least one of the nine cannabis products assessed (n=4,273).**

|  | **Monthly or more frequent use of at least one product**  **(vs. No MMF of any product)** |
| --- | --- |
| **Medical cannabis use** *(vs No medical use)* |  |
| People with a prescription | **1.32 (1.24, 1.40)** |
| People without a prescription that use medically | **1.21 (1.14, 1.28)** |
| **Age** *(vs 56-65)* |  |
| 16-25 | 1.09 (0.98, 1.21) |
| 26-35 | **1.15 (1.04, 1.27)** |
| 36-45 | **1.15 (1.04, 1.27)** |
| 46-55 | 1.04 (0.93, 1.17) |
| **Sex at birth** *(vs Male)* |  |
| Female | **0.83 (0.79, 0.87)** |
| **Race/ethnicity** *(vs White)* |  |
| Asian or Asian British | 0.88 (0.78, 1.00) |
| Black, Black British, Caribbean or African | 1.07 (0.99, 1.14) |
| Mixed or multiple ethnic groups | 1.07 (0.96, 1.20) |
| Other/Unstated | 1.08 (0.89, 1.31) |
| **Highest level of education** *(vs Less than high school)* |  |
| High school diploma | 1.03 (0.95, 1.13) |
| Some college or technical vocation | 0.94 (0.87, 1.03) |
| Bachelor’s degree or higher | 0.98 (0.90, 1.06) |
| **Income adequacy** *(vs Very/Difficult)* |  |
| Neither easy nor difficult | 1.02 (0.96, 1.08) |
| Very Easy or easy | 0.97 (0.91, 1.03) |
| Unstated | 1.13 (0.98, 1.29) |
| **Region** *(vs England no London)* |  |
| London | 1.03 (0.97, 1.09) |
| Wales | 0.98 (0.87, 1.11) |
| Scotland | 1.04 (0.96, 1.13) |
| Northern Ireland | 0.95 (0.79, 1.14) |
| **Year** *(vs 2023)* |  |
| 2024 | 1.03 (0.97, 1.08) |
| **Survey device used** *(vs Smartphone)* |  |
| Tablet | 0.99 (0.84, 1.16) |
| Computer | 0.99 (0.93, 1.04) |

Monthly or more frequent use of at least one product: A combined binary variable was created to capture the MMF use of at least one product versus no MMF use of any product. First, a continuous variable was created summing the monthly or more frequent use of each product, where 1=Yes to MMF use and 0=No to MMF use. Next, the variable was categorised to “No MMF use of any product” (0) and MMF use of at least one product (1-9).

People with a medical cannabis prescription (aRR=1.32; 1.24-1.40) and people without a medical cannabis prescription that used medically (aRR=1.21; 1.14-1.28) had a higher probability of MMF use of at least one product, after adjusting for covariates, than people that did not report medical use.

**Supplemental Table 3: Univariable and multivariable regression analyses on the monthly or more frequent use of nine cannabis products and the use of cannabis from legal medical prescription (n=4,206).**

|  | **Monthly or more frequent use of…** (vs Other) | | | | | | | | |
| --- | --- | --- | --- | --- | --- | --- | --- | --- | --- |
|  | **Dried flower** | **Oil or liquid drops** | **Oil or liquid capsules** | **Vape oils** | **Edibles** | **Cannabis drinks** | **Solid concentrates** | **Hash or kief** | **Topicals** |
| **‘All’ of their cannabis that they used in the past year was from a legal medical prescription**  (vs. Other) | RR (95% CI) | RR (95% CI) | RR (95% CI) | RR (95% CI) | RR (95% CI) | RR (95% CI) | RR (95% CI) | RR (95% CI) | RR (95% CI) |
|  | **0.70 (0.54, 0.91)** | **2.40 (1.69, 3.40)** | **1.66 (1.07, 2.56)** | **1.38 (1.00, 1.89)** | 1.25 (0.87, 1.80) | 1.39 (0.89, 2.18) | **1.63 (1.04, 2.55)** | 0.98 (0.57, 1.70) | **2.13 (1.34, 3.40)** |
|  | aRR (95% CI) | aRR (95% CI) | aRR (95% CI) | aRR (95% CI) | aRR (95% CI) | aRR (95% CI) | aRR (95% CI) | aRR (95% CI) | aRR (95% CI) |
|  | **0.72 (0.57, 0.93)** | **2.12 (1.52, 2.98)** | 1.44 (0.94, 2.21) | 1.31 (0.94, 1.82) | 1.22 (0.86, 1.74) | 1.31 (0.85, 2.01) | **1.54 (1.01, 2.36)** | 0.99 (0.58, 1.70) | **1.92 (1.22, 3.00)** |

People who reported that ‘all’ their cannabis they had used in the past year was from a legal medical prescription had a lower probability of MMF use of dried flower (aRR= 0.72; 0.57-0.93) and a higher probability of MMF use of oils or liquid drops (aRR=2.12; 1.52-2.98), solid concentrates (aRR=1.54; 1.01-2.35) and topicals (aRR=1.92; 1.22-3.00), after adjusting for covariates.

**Supplemental Table 4: Multivariable regression analyses on the characteristics of people who have ever asked for a medical cannabis prescription and who have ever received a medical prescription for cannabis in the past year (n=4,325).**

|  | **Ever asked a licensed healthcare professional for a prescription to use medical cannabis**  **(vs. No)** | **Received a prescription to use medical cannabis in the past year**  **(vs. No)** |
| --- | --- | --- |
| **High-risk use via CUDIT-SF** *(vs negative screen)* |  |  |
| Positive screen | **2.54 (2.15, 3.01)** | **3.09 (2.51, 3.79)** |
| **Age** *(vs 56-65)* |  |  |
| 16-25 | 1.44 (0.95, 2.19) | 1.89 (0.98, 3.64) |
| 26-35 | **1.52 (1.01, 2.29)** | **2.36 (1.25, 4.47)** |
| 36-45 | **1.63 (1.09, 2.24)** | **2.52 (1.34, 4.74)** |
| 46-55 | 0.96 (0.60, 1.53) | 1.25 (0.61, 2.56) |
| **Sex at birth** *(vs Male)* |  |  |
| Female | **0.76 (0.65, 0.88)** | **0.79 (0.66, 0.95)** |
| **Race/ethnicity** *(vs White)* |  |  |
| Asian or Asian British | 1.03 (0.74, 1.42) | 1.15 (0.77, 1.72) |
| Black, Black British, Caribbean or African | 1.12 (0.91, 1.37) | 1.06 (0.84, 1.35) |
| Mixed or multiple ethnic groups | 1.03 (0.77, 1.37) | 1.11 (0.79, 1.55) |
| Other/Unstated | 1.26 (0.72, 2.22) | 0.92 (0.41, 2.06) |
| **Highest level of education** *(vs Less than high school)* |  |  |
| High school diploma | 1.47 (0.97, 2.21) | 1.36 (0.78, 2.34) |
| Some college or technical vocation | **1.73 (1.20, 2.50)** | **1.82 (1.12, 2.95)** |
| Bachelor’s degree or higher | **2.21 (1.54, 3.15)** | **2.36 (1.49, 3.74)** |
| **Income adequacy** *(vs Very/Difficult)* |  |  |
| Neither easy nor difficult | 0.69 (0.54, 0.87) | 1.01 (0.75, 1.36) |
| Very Easy or easy | 1.16 (0.97, 1.42) | 1.48 (1.15, 1.91) |
| Unstated | 0.25 (0.07, 0.87) | 0.93 (0.35, 2.42) |
| **Region** *(vs England no London)* |  |  |
| London | **1.33 (1.15, 1.53)** | **1.42 (1.19, 1.69)** |
| Wales | 1.11 (0.79, 1.56) | 0.85 (0.52, 1.38) |
| Scotland | 1.16 (0.84, 1.60) | 1.14 (0.74, 1.76) |
| Northern Ireland | 1.09 (0.61, 1.93) | 1.00 (0.47, 2.15) |
| **Year** *(vs 2023)* |  |  |
| 2024 | 1.19 (0.99, 1.40) | 1.12 (0.92, 1.37) |
| **Survey device used** *(vs Smartphone)* |  |  |
| Tablet | 0.90 (0.56, 1.46) | 0.51 (0.25, 1.05) |
| Computer | 1.17 (0.99, 1.37) | 1.19 (0.97, 1.45) |
